# Outcomes among HIV-positive patients hospitalized with COVID-19

**DOI:** 10.1101/2020.05.07.20094797

**Authors:** Savannah Karmen-Tuohy, Philip M. Carlucci, Ioannis M. Zacharioudakis, Fainareti N. Zervou, Gabriel Rebick, Elizabeth Klein, Jenna Reich, Simon Jones, Joseph Rahimian

**Author notes:** These authors contributed equally and share first authorship (Savannah Karmen-Tuohy, Philip M. Carlucci). **<u>Corresponding author:</u>** Joseph Rahimian, MD, NYU Grossman School of Medicine, Department of Medicine, 31 Washington Square West, Floor number 4, New York, NY 10011.

## Abstract

**Background:** SARS-CoV-2 infection continues to cause significant morbidity and mortality worldwide. Preliminary data on SARS-CoV-2 infection suggests that some immunocompromised hosts experience worse outcomes. We performed a retrospective matched cohort study to characterize outcomes in HIV-positive patients with SARS-CoV-2 infection.

**Methods:** Leveraging data collected from electronic medical records for all patients hospitalized at NYU Langone Health with COVID-19 between March 2, 2020 and April 23, 2020, we matched 21 HIV-positive patients to 42 non-HIV patients using a greedy nearest neighbor algorithm. Admission characteristics, laboratory results, and hospital outcomes were recorded and compared between the two groups.

**Results:** While there was a trend toward increased rates of ICU admission, mechanical ventilation, and mortality in HIV-positive patients, these differences were not statistically significant. Rates for these outcomes in our cohort are similar to those previously published for all patients hospitalized with COVID-19. HIV-positive patients had significantly higher admission and peak CRP values. Other inflammatory markers did not differ significantly between groups, though HIV-positive patients tended to have higher peak values during their clinical course. Three HIV-positive patients had superimposed bacterial pneumonia with positive sputum cultures, and all three patients expired during hospitalization. There was no difference in frequency of thrombotic events or myocardial infarction between these groups.

**Conclusion:** This study provides evidence that HIV coinfection does not significantly impact presentation, hospital course, or outcomes of patients infected with SARS-CoV-2, when compared to matched non-HIV patients. A larger study is required to determine if the trends we observed apply to all HIV-positive patients.

## INTRODUCTION

The coronavirus disease of 2019 (COVID-19) pandemic, caused by the novel severe acute respiratory syndrome coronavirus 2 (SARS-CoV-2), has resulted in significant morbidity and mortality around the world. Since first tracking the global outbreak in December 2019, researchers have reported worse outcomes for patients with pre-existing conditions, including diabetes, hypertension, cardiovascular disease, underlying respiratory disease, and cancer^1,2^. However, there is minimal data exploring the effect of SARS-CoV-2 infection on the world’s estimated 37.9 million HIV-positive patients^3^. A single clinical case series of five patients and one case report have described the characteristics and outcomes of HIV-positive patients, but, to our knowledge, nothing has been published to date comparing a cohort of HIV-positive patients to a matched non-HIV cohort^4,5^. This retrospective observational study aims to understand whether coinfection with HIV alters the initial presentation, hospital course, and outcomes of patients infected with SARS-CoV-2.

## METHODS

Data were collected from electronic medical records (Epic Systems, Verona, WI) for all patients hospitalized with COVID-19 at any of four acute care NYU Langone Health hospitals in New York City between March 2, 2020 and April 23, 2020. Patients were included in the study if they had at least one positive COVID-19 test, were admitted to the hospital, and had either been discharged from the hospital, transitioned to hospice, or expired at time of analysis. Patients who did not test positive for COVID-19, who were never admitted to the hospital, and who had not yet completed their clinical course were excluded from the study. With an automated approach, we collected demographics, past medical history, admission vitals and laboratory results, and hospital outcomes. Upon generating our matched patients, manual chart review was performed to collect information, such as: CD4 counts, HIV medications, peak laboratory results, culture results, thrombotic events, and imaging results.

### Statistical Analysis

We identified 21 HIV-positive patients and 2,617 non-HIV patients who met the inclusion criteria. Greedy 1:2 nearest neighbor matching was employed using the MatchIt package, Version 3.0.2, in RStudio, Version 1.2.5042, to generate 42 matched non-HIV patients for our comparison group^6^. Patients were matched by admission date, age, body mass index (BMI), gender, tobacco history, and a history of chronic kidney disease, hypertension, asthma, chronic obstructive pulmonary disease, and heart failure. Descriptive statistics are presented as mean and standard deviation or median and interquartile range for continuous variables and frequencies for categorical variables. Normality of distribution for continuous variables was assessed by measures of skewness and kurtosis. A 2-tailed Student’s t test was used for parametric analysis, and a Mann Whitney U test was used for nonparametric data analysis. Pearson’s chi-squared test was used to compare categorical characteristics. Logistic regression was used to test associations among variables. All analyses were performed using STATA/SE 16.0 software (STATA Corp.).

### Study approval

The study was approved by the NYU Grossman School of Medicine Institutional Review Board. A waiver of informed consent and a waiver of the Health Information Portability Privacy act were granted.

## RESULTS

HIV-positive patients (N = 21) and matched non-HIV patients (N = 42) did not differ significantly in age, sex, race, tobacco use, or past medical history (Supplementary Table 1). In the HIV-positive cohort, 19 patients and 17 patients had CD4 count and viral load results, respectively, recorded prior to or on the first day of admission. Patients in the HIV-positive cohort had a median last CD4 count of 298/uL, and only six of the 19 patients had a last measured CD4 count less than 200/uL. 15 out of 17 patients had a viral load less than 50 copies/mL, and all patients in the HIV-positive cohort were on highly active antiretroviral therapy (HAART) prior to admission. HIV-positive and non-HIV groups did not differ statistically on initial white blood cell count, hemoglobin, absolute neutrophil count, ferritin, D-dimer, troponin, creatine phosphokinase, procalcitonin, or creatinine. HIV-positive patients had a higher absolute lymphocyte count [mean ± SD, HIV+: 1.09 ± .53 vs. non-HIV: 0.88 ± .39, p-value: 0.043] and higher C-Reactive Protein (CRP) [HIV+: 154.48 ±94.44 vs. non-HIV: 96.1 ± 90.0, p-value: 0.020] on initial laboratory results than non-HIV patients (Table 1). A greater percentage of HIV-positive patients had an abnormal finding of consolidation, infiltrate or opacity on initial chest imaging than non-HIV patients [HIV+: 19 (90.5%) vs non-HIV: 27 (64.3%)], though no statistical difference was seen between the groups when analyzing the finding of bilateral consolidation, opacity or infiltrate ever present on chest imaging during this hospitalization (Table 2).

**Table 1:** Admission characteristics among HIV-positive and non-HIV patients

|  | **HIV-positive**<br>N = 21 | **Non-HIV**<br>N = 42 | **P-value** |
| --- | --- | --- | --- |
| **Admission Characteristics** |  |  |  |
| Respiratory Rate, respirations per minute | 20 (18-25) | 20 (18-22) | 0.779 |
| Baseline Systolic BP, mmHg | 126 (115-139) | 137 (128-147) | **0.047** |
| Baseline Diastolic BP, mmHg | 74.43 ± 10.98 | 79.69 ± 12.50 | 0.053 |
| Temperature, degrees Celsius | 37.57 ± 0.76 | 37.64 ± 0.99 | 0.390 |
| White blood cell count 10<sup>3</sup>/ul | 7.2 (4.9-10.15)<br>N=20 | 6.25 (4.7-8.1)<br>N=38 | 0.365 |
| Hemoglobin, g/dL | 12.70 ± 2.19<br>N=20 | 13.48 ± 1.96<br>N=38 | 0.079 |
| Absolute neutrophil count, 10<sup>3</sup>/ul | 5.8 (3.9-8.7)<br>N=21 | 4.4 (3-6.4)<br>N=41 | 0.186 |
| Absolute lymphocyte count, 10<sup>3</sup>/ul | 1.09 ± 0.53<br>N=21 | 0.88 ± 0.39<br>N=41 | **0.043** |
| Ferritin, ng/mL | 679 (338-1446)<br>N=19 | 698 (232-1603)<br>N=34 | 0.905 |
| D-Dimer, ng/mL | 333 (210-452)<br>N=17 | 330 (202-466)<br>N=31 | 0.911 |
| Troponin, ng/mL | 0.02 (0.01-0.05)<br>N=18 | 0.015 (0.01-0.03)<br>N=33 | 0.308 |
| Creatine Phosphokinase, U/L | 239 (58-423)<br>N=14 | 161 (83-415)<br>N=23 | 0.728 |
| Procalcitonin, ng/mL | 0.2 (0.09-0.24)<br>N=19 | 0.08 (0.05-0.23)<br>N=36 | 0.095 |
| Creatinine, mg/dL | 1.14 (0.87-1.7)<br>N=20 | 1.09 (0.9-1.41)<br>N=38 | 0.830 |
| C-Reactive Protein, mg/L | 154.48 ± 94.44<br>N=18 | 96.1 ± 90.0<br>N=35 | **0.020** |
| Lactate Dehydrogenase, U/L | 449.4 ± 239.8<br>N=5 | 411.27 ± 229.6<br>N=11 | 0.383 |
| Last Measured Absolute CD4 Count, /ul | 298 (135-542)<br>N=19 | - | - |

Data are represented as median (IQR), mean ± SD, or N (%). Sample size is reported where it differed due to lab tests not performed. P-values were calculated using 2-sided t-test for parametric variables and Mann Whitney U test for nonparametric continuous variables. Pearson $\chi^2$ test was used for categorical comparisons. $P < .05$ was deemed significant. All laboratory results, except CD4 count, represent the first result found on admission.

**Table 2:** Hospital outcomes among HIV-positive and non-HIV patients

|  | **HIV-positive**<br>N=21 | **Non-HIV**<br>N=42 | **P-value** |
| --- | --- | --- | --- |
| Length of Hospital stay, days | 6 (4-13)<br>N=21 | 5 (3-10)<br>N=42 | 0.262 |
| O2 Flow rate AVG, L/min | 4.5 (2-16)<br>N=16 | 3.3 (2-12)<br>N=33 | 0.653 |
| O2 Flow rate MAX, L/min | 11 (2-38)<br>N=16 | 4 (3-15)<br>N=33 | 0.681 |
| Fraction of inspired oxygen AVG | 79.1 ± 21.4<br>N=7 | 71.88 ± 34.4<br>N=12 | 0.313 |
| Fraction of inspired oxygen MAX | 90.0 ± 17.32<br>N=7 | 81.25 ± 35.94<br>N=12 | 0.278 |
| Expired or transferred to hospice | 6 (28.6%) | 10 (23.8%) | 0.682 |
| Needed ICU | 6 (28.6%) | 7 (16.7%) | 0.271 |
| Needed Invasive Ventilation | 5 (23.8%) | 5 (11.9%) | 0.223 |
| Lactate Dehydrogenase Peak, U/L | 477.04 ± 210.37<br>N=21 | 436.30 ± 223.02<br>N=37 | 0.249 |
| C-Reactive Protein Peak, mg/L | 185.13 ± 107.35<br>N=20 | 128.06 ± 99.29<br>N=38 | **0.024** |
| Ferritin Peak, ng/mL | 1446 (493-2209)<br>N=20 | 1156 (314-2148)<br>N=36 | 0.617 |
| Procalcitonin Peak, ng/mL | 0.22 (0.11-0.42)<br>N=20 | 0.11 (0.06-0.28)<br>N=35 | 0.227 |
| White blood cell count Peak, 10<sup>3</sup>/ul | 8.3 (7.1-12.4)<br>N=21 | 7.3 (5.5-11.6)<br>N=41 | 0.125 |
| White blood cell count Low, 10<sup>3</sup>/ul | 5.1 (4.3-5.6)<br>N=21 | 4.6 (3.6-6.1)<br>N=41 | 0.828 |
| D-dimer, ng/mL |  |  | 0.315 |
|  | N=19 | N=34 |  |
| <1000 | 11 (57.9%) | 26 (76.5%) |  |
| 1000-6000 | 5 (26.3%) | 6 (17.6%) |  |
| >6000 | 3 (15.8%) | 2 (5.9%) |  |
| Abnormal initial chest x-ray* | 19 (90.5%) | 27 (64.3%) | **0.027** |
| Bilateral | 18 (94.7%) | 23 (85.2%) |  |
| Unilateral | 1 (5.3%) | 4 (14.8%) |  |
| Chest x-ray bilateral ever* | 18 (85.7%) | 29 (69.0%) | 0.152 |
| Myocardial Infarction | 1 (4.8%) | 1 (2.4%) | 0.611 |
| Pulmonary Embolism | 1 (4.8%) | 1 (2.4%) | 0.611 |
| Deep Vein Thrombosis | 1 (4.8%) | 1 (2.4%) | 0.611 |

Data are represented as median (IQR), mean ± SD, or N (%). Sample size is reported where it differed due to lab tests not performed. P-values were calculated using 2-sided t-test for parametric variables and Mann Whitney U test for nonparametric continuous variables. Pearson $\chi^2$ test was used for categorical comparisons. $P < .05$ was deemed significant. Peak laboratory results represent the highest measured value during hospitalization.
\*Chest x-rays were considered abnormal if a finding of consolidation, infiltrate, or opacity suggestive of pneumonia/pneumonitis was recorded.

While there was a trend toward HIV-positive patients experiencing longer hospital stays and higher rates of ICU admission, mechanical ventilation, and discharge to hospice or mortality, we did not find a statistically significant difference between the HIV-positive and non-HIV cohort on these measures. Supplemental oxygen characteristics, such as average or maximum O2 flow rate and FiO2, were not statistically different between these groups, though HIV-positive patients trended toward higher O2 flow rates when compared to non-HIV patients. Though not statistically significant, HIV-positive patients had a nominally higher peak LDH, ferritin, procalcitonin, and D-dimer. HIV-positive patients had statistically significant higher peak CRP values than non-HIV patients [HIV+: 185.13 ± 107.35 vs. non-HIV: 128.06 ± 99.29, p-value: 0.024]. Because HIV-positive patients had significantly higher admission and peak CRP values, we performed a logistic regression to see if CRP predicted mortality among HIV-positive patients and found no association with mortality using admission CRP (Odds ratio: 1.007, 95% CI: 0.998-1.015, N=18). However, we did observe a weakly significant association between the highest peak CRP values and mortality among HIV-positive patients (Odds ratio: 1.026, 95% CI: 1.002-1.051, N=20). A similar weak association was found among non-HIV patients (Odds ratio: 1.018, 95% CI: 1.006-1.029, N=38). Last measured CD4 count did not associate with mortality in HIV-positive patients (Odds ratio: 0.996, 95% CI: 0.992-1.11).

Both groups were evaluated for complications of stroke, myocardial infarction, and pulmonary embolism and deep vein thrombosis. No patients included in this study experienced a stroke. Two patients, one each from the HIV-positive and non-HIV groups, experienced both a pulmonary embolism and ST-segment elevation myocardial infarction documented on imaging, EKG, or clinical record.

The presence of a superimposed bacterial pneumonia was also evaluated. Twelve total patients (six HIV-positive, six non-HIV) had sputum cultures performed due to clinical suspicion of bacterial superinfection. Four patients had positive sputum cultures, three of whom were HIV-positive. Of these patients with positive sputum results, two patients had polymicrobial infections, and cultures grew *Pseudomonas aeruginosa* (2 patients), *Stenotrophomonas maltophilia* (2 patients), *Klebsiella pneumoniae* (1 patient), *Staphylococcus aureus* (1 patient), and *Escherichia coli* (1 patient). All four patients with positive cultures were subsequently treated with antibiotics for bacterial pneumonia, and all four expired in the hospital. Clinical suspicion of a superimposed pneumonia occurred at least six days or more prior to death in each case.

## DISCUSSION

The impact of HIV coinfection on the clinical course of patients with COVID-19 has yet to be fully characterized. Overall, our findings suggest that HIV status did not significantly impact clinical outcomes in patients with SARS-CoV-2 infection. However, we did detect trends suggesting that outcomes may be worse in HIV-positive patients, as a greater percentage of HIV-positive patients required ICU-level care, mechanical ventilation, or expired or were discharged to hospice, compared to the non-HIV cohort.

A previous study investigating outcomes in all patients admitted to NYU Langone Health hospitals found that 28.1% of patients required mechanical ventilation, and 18.5% were discharged to hospice or expired^7^. In our study, both cohorts had a lower rate of mechanical ventilation (23.8% of HIV-positive patients, 11.9% of non-HIV patients) and a higher rate of mortality (28.6% of HIV-positive patients, 23.8% of non-HIV patients), as compared to data from the larger study of all patients hospitalized at NYU Langone Health. Additionally, a recent study reporting outcomes among patients in New York City found a 21% mortality rate for hospitalized patients, while a study from China reported a 28% mortality rate among hospitalized patients^1,8^, contributing to a wide range of published mortality data. The mortality rate of both cohorts in this study falls within this established range, and further investigation is merited into the impact of HIV coinfection on COVID-19 outcomes.

We found an increased frequency of chest x-ray abnormalities on admission imaging in HIV-positive patients, and this trend persisted throughout the course of hospitalization. A prior study reported up to 84% of COVID-19 patients present with abnormal chest x-rays on admission, an incidence similar to the HIV-positive cohort in our study^9^. Admission x-ray was not a reliable indicator of illness severity in HIV-positive patients, a similar finding to what has been reported in the general population^9^.

In this cohort, four patients were clinically treated for superimposed bacterial infection based on positive sputum culture results, three of whom were HIV-positive. All three HIV-positive patients with bacterial superinfection expired in the hospital. Existing data highlights the higher incidence of bacterial pneumonias in HIV-positive patients compared to the general population, in addition to the significant mortality burden of non-AIDS bacterial infections in this population^10-14^. Risk factors associated with contracting severe non-AIDS bacterial infections include immunosuppression and a history of cancer and diabetes, comorbidities that have also been associated with worse prognosis in SARS-CoV-2 infection^2,8,15^. This study was not powered to conclude that superimposed bacterial infections were a predictor of mortality in HIV-positive patients with SARS-CoV-2 infection. However, all HIV-positive patients with a bacterial pneumonia expired, and this finding should be considered when making clinical decisions about these patients. Future studies should investigate whether SARS-CoV-2 infection increases the risk of secondary bacterial infection in similar patients and if bacterial pneumonia might serve as a predictor of mortality in the HIV-positive population.

All HIV-positive patients in the study were on HAART prior to hospital admission, and only one patient in the HIV-positive cohort had both a CD4 count less than 200/uL and a viral load greater than 50 copies/mL. Therefore, these findings may not apply to a population with poorly controlled HIV or AIDS. Only one patient was taking a protease inhibitor, a class of drugs under investigation for potential therapeutic benefit in COVID-19 treatment^16^. Given the lack of data in our cohort, we are unable to comment on the clinical impact of protease inhibitor use in HIV-positive patients with SARS-CoV-2 infection. This was a retrospective study that is susceptible to confounding variables, and our limited sample size prevented us from detecting small differences among groups. Larger prospective studies should examine the impact of poorly controlled HIV on the SARS-CoV-2 clinical course. Taken together, our findings are reassuring that HIV-positive patients may not experience significantly worse outcomes in SARS-CoV-2 infection, as compared to matched non-HIV patients.

## Data Availability

Individual level data are not available for this study. For inquiries, please contact the corresponding author.

## ACKNOWLEDGEMENTS

The authors thank Mark Mulligan, Christopher Petrilli, Collin Ortals, Brian Bosworth, Robert Cerfolio, Steven Chatfield, Thomas Doonan, Fritz Francois, Robert Grossman, Leora Horwitz, Juan Peralta, Katie Tobin, and Daniel Widawsky for their operational and technical support. We also thank the thousands of NYU Langone Health employees who have cared for these patients.

---

**Supplementary Table 1.**

|  | HIV+<br>N = 21 | Non-HIV<br>N = 42 | P-value |
| --- | --- | --- | --- |
| Demographics |  |  |  |
| Age, years | 60.04 ± 11.77 | 61.48 ± 20.13 | 0.383 |
| Male Sex | 19 (90.5%) | 38 (90.5%) | 1.000 |
| Race |  |  |  |
| African American | 5 (23.8%) | 4 (9.5%) | 0.065 |
| White | 8 (38.1%) | 23 (54.8%) |  |
| Asian | 0 (0%) | 6 (14.3%) |  |
| Other/Unknown | 8 (38.1%) | 9 (21.4%) |  |
| History |  |  |  |
| Tobacco use |  |  | 0.851 |
| Never or Unknown | 16 (76.2%) | 31 (73.8%) |  |
| Former | 3 (14.3%) | 5 (11.9%) |  |
| Current | 2 (9.5%) | 6 (14.3%) |  |
| Hypertension | 7 (33.3%) | 16 (38.1%) | 0.711 |
| Hyperlipidemia | 4 (19.0%) | 9 (21.4%) | 0.826 |
| Coronary Artery Disease | 1 (4.8%) | 3 (7.1%) | 0.715 |
| Heart Failure | 0 (0%) | 0 (0%) | - |
| Peripheral Vascular Disease | 1 (4.8%) | 1 (2.4%) | 0.611 |
| Asthma or COPD | 4 (19.0%) | 6 (14.3%) | 0.361 |
| Diabetes | 4 (19.0%) | 8 (19.0%) | 1.000 |
| Malignancy | 3 (14.3%) | 3 (7.1%) | 0.363 |
| Transplant (completed) | 0 (0%) | 0 (0%) | - |
| Chronic Kidney Disease | 4 (19.0%) | 7 (16.7%) | 0.814 |
| BMI kg/m² | 28.21 ± 5.37 | 28.24 ± 5.18 | 0.493 |

Data are represented as median (IQR), mean ± SD, or N (%). Sample size is reported where it differed due to lab tests not performed. P-values were calculated using 2-sided t-test for parametric variables and Mann Whitney U test for nonparametric continuous variables. Pearson $\chi^2$ test was used for categorical comparisons. $P < .05$ was deemed significant.

